# Virtual environment benefits motor dysfunction in Functional Neurological Disorder: Evidence from Virtual and Augmented Reality

**DOI:** 10.64898/2026.09.09.26362633

**Authors:** Laure von der Weid, Joaquín Penalver-Andres, Cristina Concetti, Laura Marchal-Crespo, Eliane Müller, Selma Aybek

## Abstract

**Background:** Virtual and Augmented Reality (VR/AR) are increasingly used in medicine and may offer novel interventions for Functional Neurological Disorder (FND) by targeting mechanisms of impaired motor control. However, evidence for the effect of different modalities on movement in FND is limited. Moreover, it is hypothesized that visual feedback of one’s affected arm could reinforce FND symptoms through maladaptive self-monitoring. The present study aims to compare motor performance under visual feedback of a virtual avatar versus the real arm.

**Methods:** Fifteen FND patients and eighteen healthy controls completed a dual motor reaching task (touching a virtual fruit such as orange or apple appearing in front of them) with a virtual headset across three modalities: Augmented Reality (AR), where participants saw their real arm; Virtual Reality (VR), with a virtual arm; and Virtual Reality Improved (VRI), with artificially enhanced straightness and larger collision zone with the target. Their movement speed, straightness and number of velocity peaks were calculated for each fruit catch. Additionally, the number of collisions between the eyes and the body was calculated with eye tracking technology.

**Results:** FND participants were slower and produced less smooth movements in AR compared to VR (*b* = 1.09, *z* = 8.83, *P* < .001) and VRI (*b* = 0.85, *z* = 6.91, *P* < .001), while the opposite effect was found in healthy controls. Eye tracking showed that FND patients looked more at their affected hand than unaffected hand (b = −1.46, SE = 0.34, z = −4.23, p < .001; OR = 0.23).

**Conclusions:** Virtual feedback of their limb’s avatar appears to support better motor performance, while visual feedback of their real limb appears to exacerbate motor impairments in FND patients, likely reinforcing maladaptive self-monitoring. These findings highlight the therapeutic potential of VR interventions and provide the basis for future studies examining longer protocols, clinical outcome and potential implementation in standard motor rehabilitation.

## Introduction

Functional neurological disorder (FND) is a complex and common condition thought to emerge from alterations in brain network functioning.^1^ Contemporary theories propose a multifaceted interaction of emotion processing, agency, attention, interoception and predictive processing/inference.^2^ The diagnostic process has recently been subject to change, with the exclusionary nature of the FND diagnosis now being rendered obsolete. In the contemporary context, the prevailing approach is to formulate diagnoses based on positive indicators that are specific to FND.^3^ One such sign is distractibility, whereby the severity of symptoms is reduced when attention is not focused on them.^4^ This sign is of significant interest for the advancement of our understanding of the pathophysiology of the disease, and has potential for the development of new treatment plans.

One of the central mechanisms of FND can be understood via the Bayesian Brain Model. Within this theoretical framework, the brain is conceptualised as an inference model that predicts subsequent actions based on historical feedback.^5^ As posited by the aforementioned theory, the model will be updated with new incoming sensory information, in conjunction with prior knowledge. Consequently, all sensory experiences and behavioural actions are the consequence of predictions derived from previously accumulated information. In the event of new information that does not align with the prediction, a mismatch signal is generated, known as "Prediction Error". In this scenario, either the priors are updated to align with the new information, or this new information is modified to be interpreted within the context of the model.^6^ It is theorised that FND symptoms may arise from altered prediction errors.^7^ The fundamental premise asserts that, in patients, anomalous priors will be excessively robust, compelling the movement or sensation to conform to these preconceived expectations. The deficient model will then be reinforced by the knowledge of deficient movement and/or sensation. This observation aligns with the Bayesian brain theory. It is noteworthy that, during a video-recorded task, patients diagnosed with FND allocate 66% of their time looking at their tremor, as opposed to 32% in patients with organic tremors.^8^ Another neuroimaging study demonstrated increased self-monitoring, the brain’s ability to observe, evaluate, and track one’s own actions as they occur, in individuals with FND. This was evidenced by greater activation in the ventromedial prefrontal cortex (vmPFC), ventrolateral prefrontal cortex (vlPFC), and superior posterior temporal lobe.^9^ Notably, these regions showed heightened activity when participants imagined movements with the affected limb compared to the unaffected limb. These findings suggest that models emphasizing altered self-monitoring processes hold significant potential for the development of virtual reality (VR) interventions, given their relative ease of integration.

Virtual reality (VR) immerses users in a computer-generated environment, typically through a head-mounted display, in which visual feedback about one’s own movements can be manipulated in real time. In recent decades, interest in its medical applications has grown steadily,^10^ and preliminary studies suggest it could be beneficial for individuals suffering from FND.^11^ Primarily, VR is a safe technique with very few side effects that also has the ability to replicate real-world rehabilitative interventions in a clinical setting.^12,13^ It also offers promising opportunities in terms of treatment engagement and motivation.^14^ Secondly, the implementation of a VR intervention could facilitate the precise targeting of the mechanism underlying FND, notably through the updating of the Bayesian brain model with novel priors. The construction of a virtual environment with a virtual representation of one’s body allows for the display of smoother and more adequate movements, and consequently training the brain to observe new movements. Furthermore, a virtual environment offers a great source of distractibility, as the participant is not able to observe their own symptoms.

A few studies (n = 12), mostly with a limited number of participants, showed preliminary evidence that the use of VR could have some clinical benefit on FND patients,^11^ notably on posture and dizziness.^15^ However, only five of them were focusing on various motor symptoms. One showed an improvement of body control in VR in a dual task setting (motor and attentional task), but only when the attentional demand is high.^16^

In summary, VR could be a cost-efficient, non-invasive, and widely accepted treatment plan for individuals with functional neurological disorders, as well as a novel technique to study the underlying pathophysiology. However, it remains to be proven whether these interventions are feasible and effective. The present study compares three visualization modalities: Augmented Reality (AR), which overlays virtual elements onto the real environment; standard Virtual Reality (VR); and Virtual Reality Improved (VRI), in which the visual feedback of the participant’s movement is artificially enhanced. It is hypothesised that FND patients would exhibit straighter, smoother and faster movements in VR modalities when their affected limb is not visible. We also expect the performance to be improved by a parallel cognitive task due to distractibility in FND. Finally, we expect FND patients to look more at their body or avatar during the task than healthy controls.

## Materials and methods

Participants with Functional Neurological Disorder (FND) and healthy controls completed a series of motor tasks within three visualization modalities: Augmented Reality (AR), Virtual Reality (VR), and a VR condition with an artificial enhancement of performance (VRI). The order of modalities was counterbalanced across participants. Each participant also performed a physical reaching task without the head mounted display and completed questionnaires assessing their sense of control and agency across all modalities.

### Participants

18 FND participants were recruited from the Psychosomatic Medicine Unit at the Bern University Hospital (Inselspital Bern). They were recommended to take part in the study by a board-certified neurologist according to DSM-5 criteria and positive signs. Eligible symptoms comprise motor symptoms (F44.4; abnormal movements and weakness) in the upper arms (left, right or both arms). Participants who additionally experienced functional seizures (F44.5), sensory deficit (F44.6) and mixed symptoms (F44.7) were also included. Concerning the control group, 20 age-gender-matched healthy subjects participated in the study. Healthy controls were recruited through advertisements in public media (e.g., local newspapers, university notice boards, local science dissemination events). Exclusion criteria included the presence of major comorbid psychiatric disorders, history of brain surgery, history of alcohol or drug abuse and cybersickness (i.e., nausea when looking at a screen or playing computer games). After the experiment, 3 FND participants and 2 healthy controls were excluded from the analysis due to technical errors during the experiment.

The participants signed a written informed consent form and were not paid for their participation. The protocol was approved by the local ethics committee of the Canton of Bern, Switzerland (BASEC number: 2020-02283), and was conducted in accordance with the Declaration of Helsinki.

### Clinical Data

The severity of the symptoms has been rated by the experimenter using the Clinical Global Impression Score (CGI)^17^ and self-rated by the participant on a visual analogue scale from 1 to 100. The type of symptoms, as well as the duration of the symptoms, are reported.

### Experimental Design

The participants were seated comfortably in an office chair, set at a predefined fixed location in front of the computer. The Head-Mounted Display (HMD) device was then fitted to the participant.

The HMD integrated an HTC Vive Pro Eye (HTC Vive, Taiwan) and the AR add-on Zed Mini stereo camera (StereoLabs, USA) on the HMD’s front. The HTC Vive allowed visualization of the virtual environment from a first-person perspective and tracking participants’ eye movements. Although the HTC Pro Eye also incorporates video-see-through AR capabilities, we opted for the faster ZedMini to mimic how participants visually perceive the real world by recording a high-definition, low-latency stereo video feed of the environment ahead of the HMD. Participants’s arm movements were tracked using two trackers (HTC Vive Trackers) placed on the elbow of the participant and 2 hand-held HTC Vive controllers. The virtual environment and task were created by the research team using the game engine Unity 3D (version 5.5.3).

#### Visualization modalities

The experiment consisted of three different virtual interaction modalities which all worked on the same principle: Augmented Reality (AR), i.e. participants could see their own body; Virtual Reality (VR), i.e. participants saw a virtual representation of themselves (an avatar) which moved synchronously with their arm movements; and Virtual Reality Improved (VRI), i.e. participants saw an avatar whose movements were artificially enhanced to appear smoother, without having a direct influence on the physical movement. The object’s collision zone was additionally increased. Those three modalities allowed us to study the effect of body visibility on self-generated movement performance (**Fig 1).**

**Figure 1.**
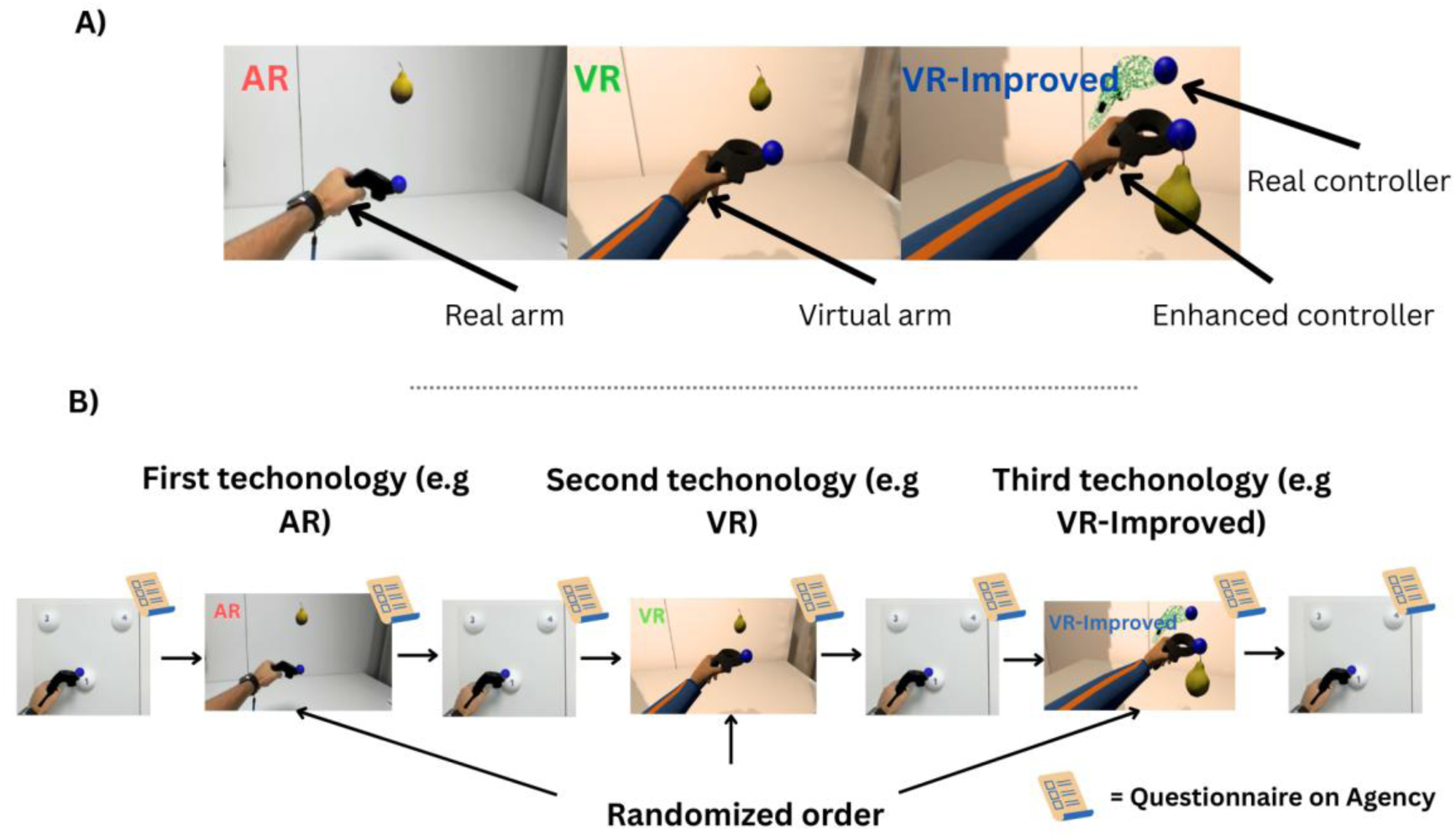
Study protocol and experimental modalities. **A)** Illustration of the three movement modalities: Augmented Reality (AR), in which participants see their own physical arm; Virtual Reality (VR), in which their arm is replaced by a virtual limb; and Virtual Reality Improved (VR-Improved), in which the virtual movement is artificially enhanced to appear smoother and more efficient. **B)** Overview of the experimental sequence. All participants first performed a physical reaching task, followed by one of the virtual modalities. They then alternated between virtual modalities and additional physical reaching blocks. After every movement task, participants filled out the questionnaires. During each task, the experimenter asked regularly to change hands.

### Virtual task

The virtual task was based on the one developed by Wenk et al.^18^ The reader is referred to this work for more detailed information. It consisted of a dual motor-cognitive task where participants reached virtual collectibles (fruits [apples, oranges and pears]; i.e., motor task) visually presented in front of them using the HMD while keeping the count of different types of collectibles caught (types of fruits; i.e., cognitive task). The fruits appeared at 22 pre-programmed locations in 3D space, in a counterbalanced order and fruit-location assignment.

Participants performed the task sequentially with both arms in counterbalanced order, so that limb dominance and, in patients, limb involvement were balanced across conditions rather than confounded with them. Finally, participants also performed the motor task alone (i.e., without counting the types of fruit) to examine the effects of distractibility on motor performance during the task.

At any given moment, only one fruit was presented on the screen. Participants reached/caught the fruit by moving their hand from a green sphere virtually presented in the center of the movement space towards the fruit, making contact with the controller. Once “touched’’, the fruit disappeared and the green sphere re-appeared in the center of the workspace. Participants were instructed to stay on the green sphere with the controller and hold contact until it disappeared, after which the next fruit appeared (unless the session had ended).

All participants took part in all modalities. Each modality started and ended with a test session, consisting of 6 reaching movements with the left hand and 6 with the right hand (order counterbalanced across participants), which only contained pears and without the cognitive task. Those test sessions result in two time points “*before*” and “*after*”. Between those test sessions, participants performed a *task phase* with 18 movements with the left hand and 18 movements with the right hand while counting fruits (motor and cognitive task), as well as 18 movements with each hand without counting the fruits (only motor task). This led to 12 movements for each test (24 in total with the two tests) and 72 movements for the task part. Overall, each modality comprised 96 reaching movements. Once all the trials of one modality were done, participants were directly asked to fill out questionnaires to rate their performance and their agency over the self-generated movements and agency/ownership.

### Physical reaching

To evaluate a potential effect of the task beyond the virtual modalities, participants were also asked to reproduce the movement by reaching and touching polystyrene balls. This consisted of three balls that the participant had to physically touch with the controller, which also recorded the movement. The balls had to be touched by both the right and left hands, once in a clockwise direction and once in a counterclockwise direction, leading to 12 movements in total (3 balls touching clockwise with the left and right arms, 3 balls touching counterclockwise with the left and right arms).

### Questionnaire

At the end of each virtual modality (AR, VR, VRI) and the physical reaching, participants were asked to rate their performance (judgement of performance – JoP) and their sense of control over the virtual/real hand (judgement of agency – JoA). Additionally, after each virtual modality (AR, VR and VRI), they filled out 10 questions on their sense of Ownership, Agency, Control and Dis-ownership **(See supplementary material)**. The responses were noted on a 7-point Likert Scale. All questionnaires were filled out on paper.

### Protocol

The study used a within-subject design in which all participants (HC x FND) completed all three virtual modalities. Each session began with a physical reaching block, after which the three virtual modalities (AR x VR x VRI) were administered in a counterbalanced order across participants, with a physical reaching block interleaved between modalities and a final one at the end. Participants completed the questionnaire after every physical reaching block and every virtual modality. **Fig 1**.

### Data processing and analysis: Arm movements

Data processing and analysis were performed in Python (version 3.12.8) and RStudio (version 4.3.2). The movement data recorded from the handheld controller were segmented into individual fruit-reaching trials. Each trial began when the green sphere disappeared, revealing the fruit, and ended when the fruit disappeared, indicating contact between the controller held by the hand representation and the fruit.

To assess movement quality, several kinematic outcomes were extracted from the controller’s trajectory: (1) normalised movement duration (s/m), calculated as the total movement time divided by the minimum distance between the start and end positions; (2) trajectory straightness ratio, defined as the actual path length divided by the minimum straight-line distance between the start and end positions; (3) the number of velocity peaks, which corresponds local maxima of the speed profile whose prominence exceeded 5% of that movement’s peak speed, reflecting the movement smoothness.^18^ Normalized movement duration indexes the time taken to reach the target, accounting for the target’s distance, thus informing overall movement speed. Movement straightness informs on how the actual trajectory differs from the theoretical, minimal-distance trajectory, therefore informing on the efficacy of the movement.

To filter for eventual outlier data points due to technical issues and extreme data, all participants’ individual medians were computed for each outcome. Then, every trial that was farther than 3 median absolute deviation (MAD) from the participant’s median was deleted. In total, 7.11% of trials were suppressed in the FND group and 6.68% in the HC group. Participants were additionally screened for outliers, defined as values exceeding ±3 SD from the mean, on each outcome within each modality. As no participant showed consistently outlying values across modalities and outcomes, none was excluded from the analyses.

For analyses involving between-group factors, Pearson’s Chi-squared test, Welch’s t-test, or Wilcoxon rank-sum tests were applied as appropriate. To account for repeated measures within participants, linear mixed-effects models (LMMs) were conducted for each outcome using the lme4 package in Rstudio, with *ParticipantType* (FND, HC), *Modality* (AR, VR, VRI), *CognTask* (Count, No-count), or *test time point* (before the task phase and after the task phase) as fixed factors, and participant (*Pcode*) as a random intercept. P values for the coefficient significance tests were estimated using the ‘lmerTest’ package (Kuznetsova et al., 2017) and contrasts were obtained with the ‘emmeans’ package. No covariates were included in the models. Modality comparisons were Tukey-corrected within the group; group comparisons were Holm-corrected across modalities.

Concerning the *test time point* analysis (the 12 first movements of a modality vs the 12 last), only FND patients have been analyzed to decrease the complexity of analysis and as the intervention is more relevant to the clinical population.

During the counting trials, participants reported the number of fruits they had touched. A trial was scored as correct when the reported number matched the number touched, and as incorrect otherwise. When a participant made consecutive counting errors, only the first was scored as incorrect; subsequent trials within the same error sequence were scored as correct, as their accuracy was determined by the preceding error rather than by an independent counting failure. Counting accuracy was analysed at the trial level using a generalized linear mixed-effects model (GLMM) with a binomial distribution and logit link. Group (FND vs. HC) was entered as a fixed effect, and a by-participant random intercept accounted for the repeated trials within each participant.

### Data processing and analysis: Eye tracking

Four target regions were defined for gaze classification: the participant’s left hand, left arm, right hand, and right arm. All remaining visual space, including the fruit used during the task, was categorized as the environment. For each trial, gaze samples were mapped onto these predefined regions, and a binary trial-level variable was generated for each target: a trial (i.e., a single arm movement) was coded 1 for a given target if any fixation fell within that target’s region at any point during the trial, and 0 otherwise. Because participants fixated the environment in 100% of trials, this category was excluded from subsequent analyses.

For the main analysis, trial-level binary gaze outcomes were analyzed using generalized linear mixed-effects models (GLMMs) with a logit link. Group (FND vs. HC) was entered as a between-subjects fixed effect and participants as random intercepts, to test whether FND participants oriented their gaze toward their own body more frequently than HC. Because body-directed gaze occurred almost exclusively in the AR condition, with too few observations in VR and VRI to support estimation, this analysis was restricted to AR.

A secondary analysis, within the FND group only and in the AR condition, compared body-directed gaze between trials performed with the affected versus the unaffected hand, using the same GLMM framework, to determine whether gaze allocation differed according to the symptom-affected limb.

### Data analysis: Questionnaire

For each questionnaire, item responses belonging to the same construct (e.g., ownership, agency, etc.) were averaged to obtain a single score per category for each participant and modality. Because this procedure resulted in one summary score per participant per modality, questionnaire outcomes were analyzed using a two-way mixed ANOVA with Group (FND vs. HC) as a between-subjects factor and Modality (AR, VR, VRI) as a within-subjects factor. This analysis tested whether subjective experiences (e.g., ownership and agency) differed between groups and across modalities. Results can be found in the supplementary materials.

## Results

### Clinical and Demographic Data

The two groups did not differ in age or sex (P = 0.64 and P = 0.87, respectively), nor in their use of psychotropic medication (P = 0.065). Among the FND participants, 8 had the right arm affected, 3 had the left arm affected, and 3 had bilateral arm symptoms. **Table 1**.

**Table 1:**
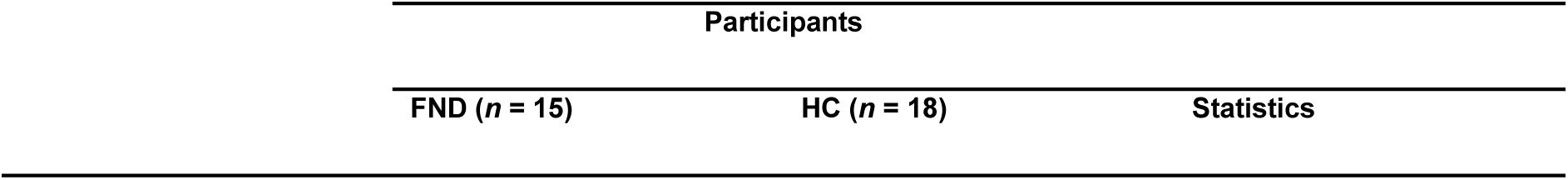
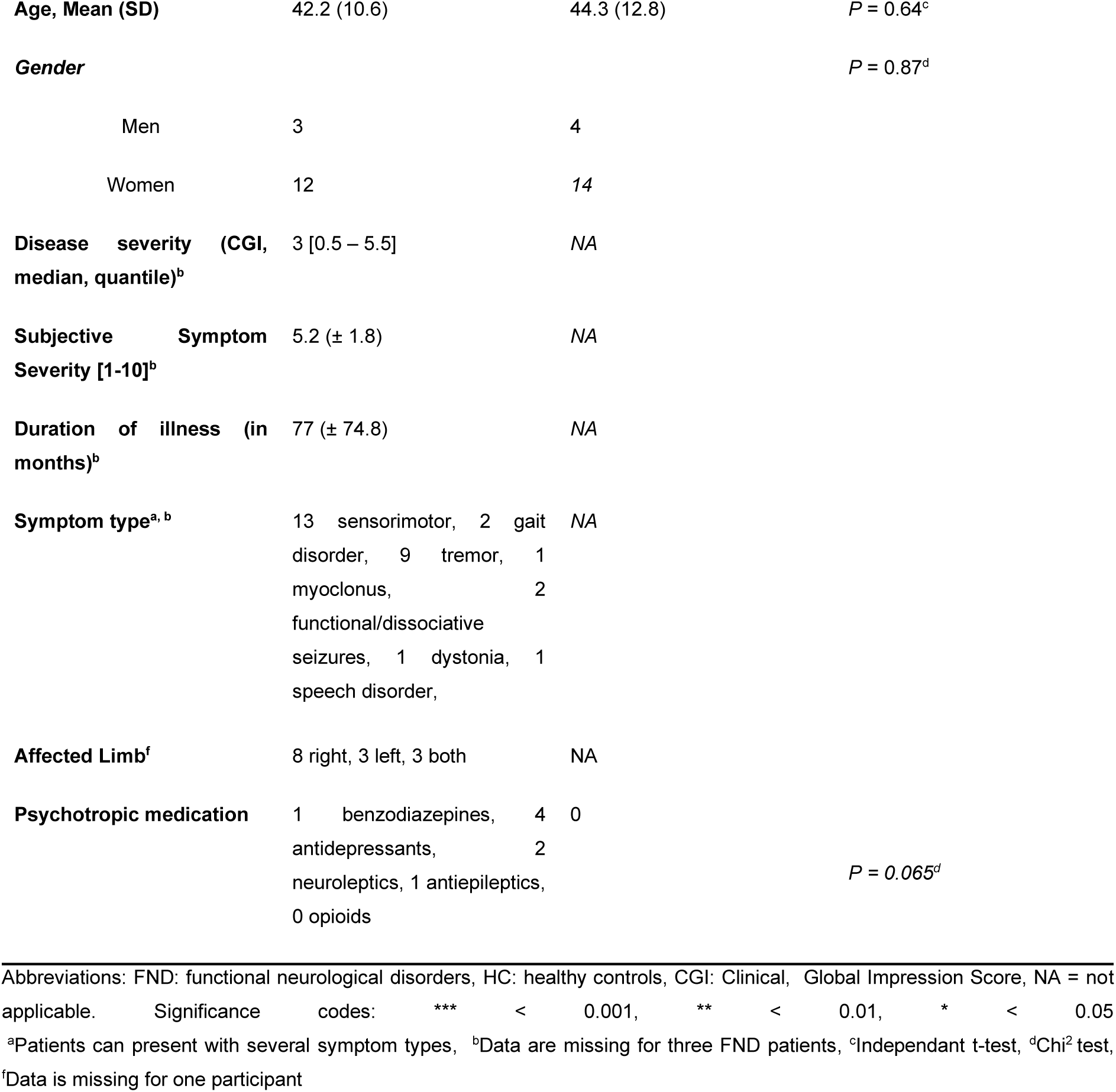
Summary of demographic and clinical characteristic in both groups.

### FND patients perform reaching movements more slowly and less smoothly than HC, especially when seeing their own arm

Normalized movement duration showed a significant Group × Modality interaction (F(2, 3429) = 42.16, P < .001), together with main effects of Modality (F(2, 3429) = 17.29, P < .001) and Group (F(1, 31) = 5.01, P = .033). Because the interaction was significant, group differences were examined within each modality. This difference was present in the AR modality (b = 2.97, SE = 0.85, z = 3.51, P < .001, P_Holm_ = .002). In the VRI modality, it did not survive multiple comparison (b = 1.89, SE = 0.85, z = 2.23, P = .026, P_Holm_ = .051), and was absent in VR (b = 1.37, SE = 0.85, z = 1.62, P = .106). Within the FND group, normalized movement duration was significantly higher in AR compared to VR (b = 1.09, SE = 0.12, z = 8.83, P < .001, P_Tukey_ < .001) and compared to VRI (b = 0.85, SE = 0.12, z = 6.91, P < .001, P_Tukey_ < .001), with no significant difference between VR and VRI (b = –0.24, SE = 0.12, z = –1.97, P = .12). Healthy controls showed the reverse pattern, moving faster in AR than VR (b = –0.52, SE = 0.11, z = – 4.56, P < .001, P_Tukey_ < .001), although the difference between AR and VRI was not significant (b = –0.23, SE = 0.11, z = –2.09, P = .092). They were also faster in VRI than VR (b = 0.28, SE = 0.11, z = 2.54, P = .03, P_Tukey_ = .03). **Graph 1.**

**Graph 1.**
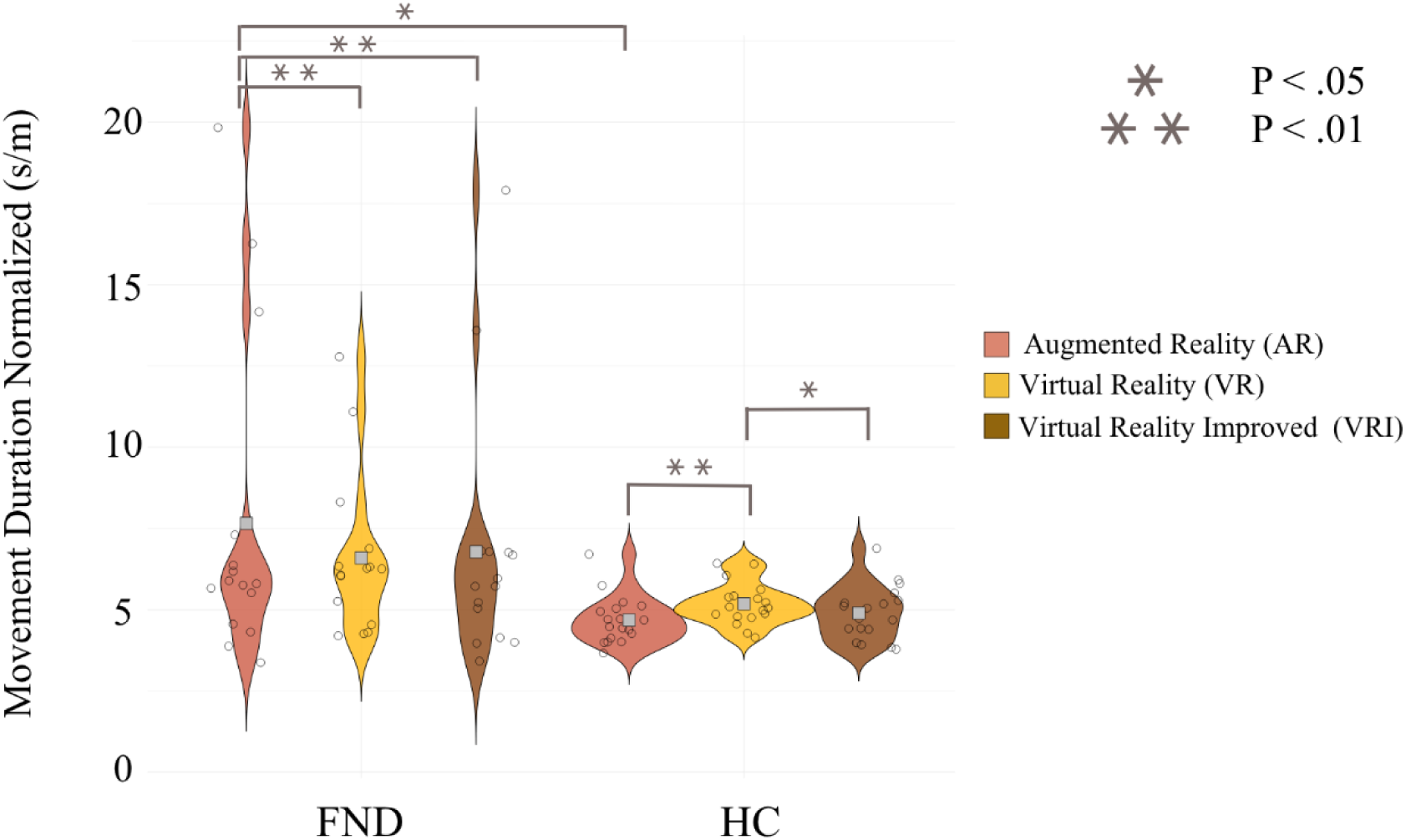
Mean movement duration across groups and modalities. FND participants are slower in the AR modality, whereas healthy controls are faster in this modality compared to the other.

For the number of Peaks, we observed a Modality effect (F(2, 3429) = 30.11, P < .001), a group effect with FND patients producing significantly more velocity peaks than healthy controls (F(1, 31) = 5.62, P = .024) and a positive interaction (F(2, 3429) = 13.67, P < .001). The simple effect showed that FND produced more peaks only in the AR condition (b = 1.86, SE = 0.398, z = 4.66, P < .001, P_Holm_ < .001). This group difference was not observed in VR (b = 0.637, SE = 0.398, z = 1.59, P = .109) or VRI (b = 0.756, SE = 0.398, z = 1.89, P = .058). Within the

FND group, AR again elicited the greatest number of peaks, with significantly more peaks than both VR (b = 1.186, SE = 0.116, z = 10.17, P < .001, P_Tukey_ < .001) and VRI (b = 1.137, SE = 0.116, z = 9.74, P < .001), while VR and VRI did not differ (b = −0.049, SE = 0.116, z = −0.42, P = .907). In healthy controls, none of the contrasts were significant (AR–VR: b = −0.036, SE = 0.106, z = −0.34, P = .938; AR–VRI: b = 0.033, SE = 0.106, z = 0.311, P = .947; VR–VRI: b = 0.069, SE = 0.106, z = 0.65, P = .791), indicating comparable movement smoothness across modalities. **Graph 2.**

**Graph 2.**
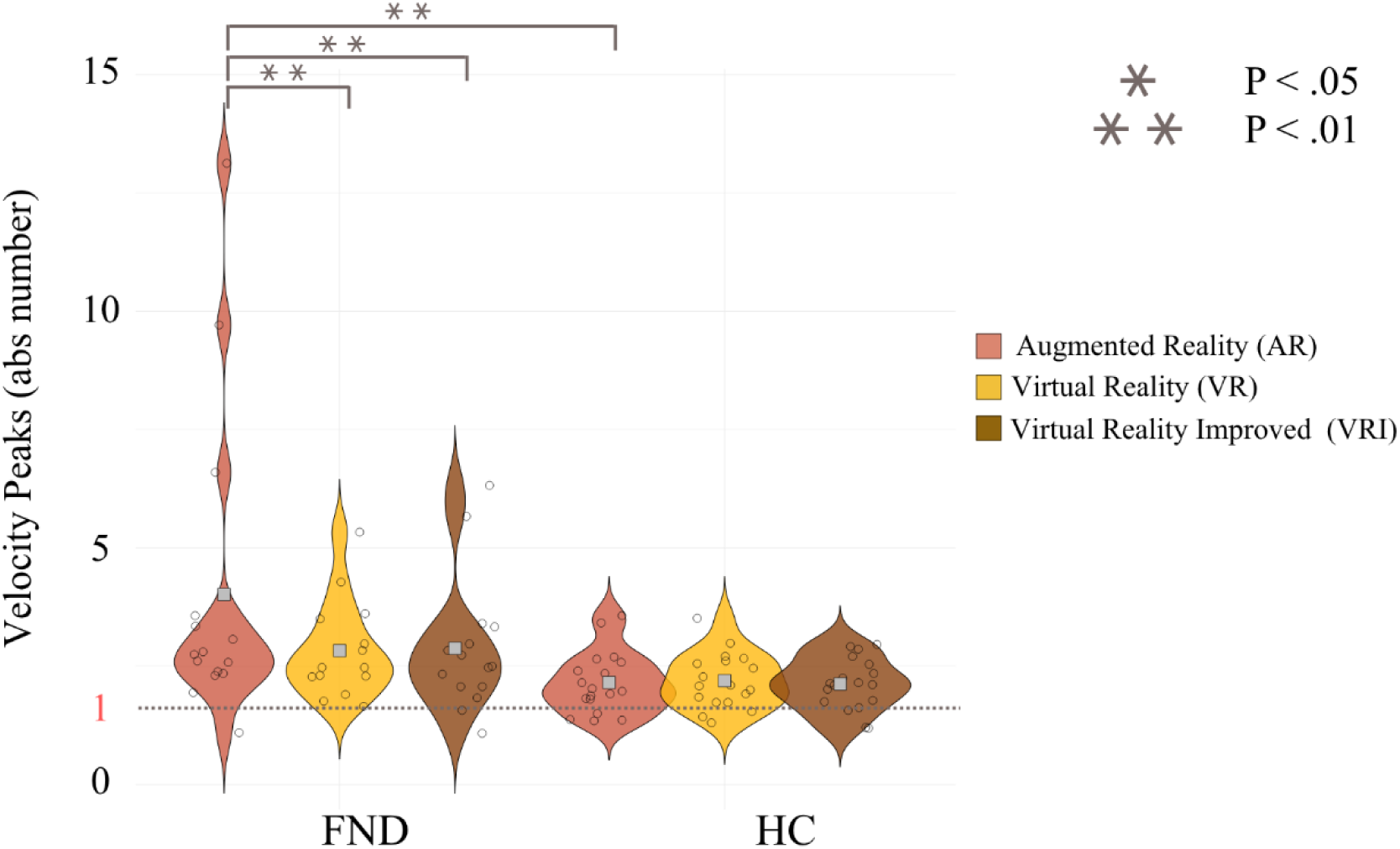
Mean number of peaks across groups and modalities. FND patients produce more peaks in the AR condition compared to VR and VRI as well as compared to HC in AR. HC produces a number of peaks closer to 1, which is expected in this task.

For movement straightness, there was no overall difference between FND patients and healthy controls (F(1, 31) = 2.09, P = .15), no interaction (F(2, 3429) = 0.09, P = .91) but a modality effect (F(2, 3429) = 12.55, P < .001). The absence of group difference was consistent across modalities (AR: b = 0.09, SE = 0.064, z = 1.41, P = .157; VR: b = 0.07, SE = 0.064, z = 1.14, P = .253; VRI: b = 0.08, SE = 0.063, z = 1.29, P = .19). Within groups, both FND patients and healthy controls showed straighter movements in VRI than in either AR or VR. For FND, straightness was higher in VRI compared to AR (b = 0.051, SE = 0.011, z = 4.56, P < .001, P_Tukey_ < .001) and compared to VR (b = 0.034, SE = 0.011, z = 3.05, P = .006, P_Tukey_ = .006), with no difference between AR and VR (b = 0.017, SE = 0.01, z = 1.55, P = .266). Healthy controls showed the same pattern, with straighter movements in VRI compared to AR (b = 0.044, SE = 0.01, z = 4.22, P < .001, P_Tukey_ < .001) and compared to VR (b = 0.043, SE = 0.01, z = 4.23, P < .001, P_Tukey_ = .006), and again no difference between AR and VR (b = 0.0001, SE = 0.01, z = 0.014, P = .999). **Graph 3.**

**Graph 3.**
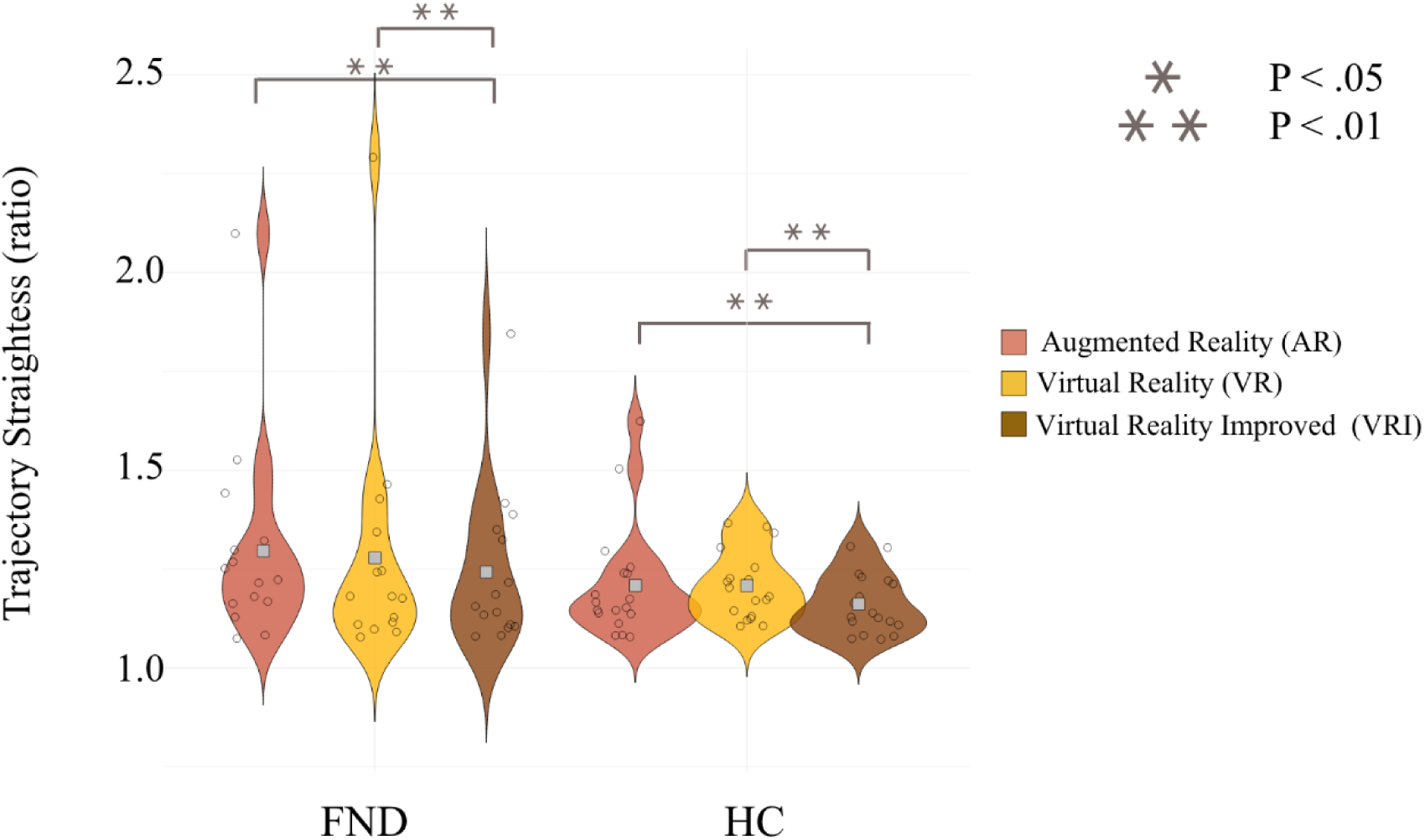
Mean straightness across groups and conditions. Both groups show a more straight movement (closer to 1) in the VRI modality compared to AR and VR. No group difference was observed.

**Table 2:**
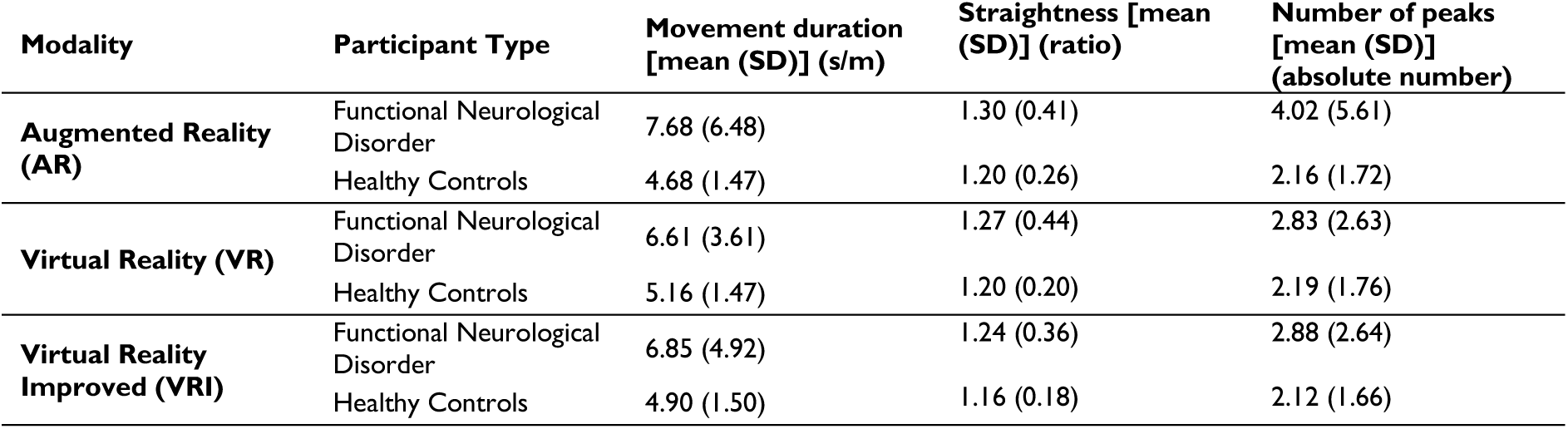
Mean of each outcome by group and modality during the task.

| Modality | Participant Type | Movement duration [mean (SD)] (s/m) | Straightness [mean (SD)] (ratio) | Number of peaks [mean (SD)] (absolute number) |
| --- | --- | --- | --- | --- |
| **Augmented Reality (AR)** | Functional Neurological Disorder | 7.68 (6.48) | 1.30 (0.41) | 4.02 (5.61) |
|  | Healthy Controls | 4.68 (1.47) | 1.20 (0.26) | 2.16 (1.72) |
| **Virtual Reality (VR)** | Functional Neurological Disorder | 6.61 (3.61) | 1.27 (0.44) | 2.83 (2.63) |
|  | Healthy Controls | 5.16 (1.47) | 1.20 (0.20) | 2.19 (1.76) |
| **Virtual Reality Improved (VRI)** | Functional Neurological Disorder | 6.85 (4.92) | 1.24 (0.36) | 2.88 (2.64) |
|  | Healthy Controls | 4.90 (1.50) | 1.16 (0.18) | 2.12 (1.66) |

### FND patients are more impacted by the cognitive task

During half of the trials, participants were asked to count the number of each type of fruit they have catched (number of oranges, apples). Comparing the effect of the cognitive task, both groups were slower while counting (F(1, 6517) = 391.83, P < .001), and FND were slower than controls (F(1, 31) = 6.02, P = .019). This difference was found in both the No-Count condition (b = 1.90, SE = 0.84, z = 2.26, P = .023, P_Holm_ = .025) and the Count condition (b = 2.26, SE = 0.84, z = 2.65, P = .007, P_Holm_ = .016). Within-group contrasts showed that both groups were slower when counting (FND: b = –1.48, SE = 0.097, z = –15.14, P_Tukey_ < .001; HC: b = –1.14, SE = 0.086, z = –12.77, P_Tukey_ < .001). The interaction indicated that FND participants were more affected by the cognitive task than HC (F(2, 6517) = 6.49, P = .01). For movement straightness, both groups showed less straight trajectories in the Count condition (F(2, 6517) = 266.89, P < .001) (FND: b = –0.130, SE = 0.009, z = –14.31, P_Tukey_ < .001; HC: b = –0.072, SE = 0.008, z = –8.60, P_Tukey_ < .001), with a significant interaction showing stronger deterioration in FND (F(2, 6517) = 21.64, P < .001). The number of peaks showed the same pattern: both groups produced more peaks in the Count condition (F(2, 6517) = 7.79, P = .009) (FND: b = – 0.94, SE = 0.095, z = –9.89, P_Tukey_ < .001; HC: b = –0.64, SE = 0.087, z = –7.35, P_Tukey_ < .001), and the interaction confirmed a greater effect of the cognitive task in FND (F(2, 6517) = 5.49, P = 0.019). **Graph 4.**

Counting accuracy did not differ significantly between groups (FND: 78.8%, HC: 75.6% of trials counted correctly; OR = 0.83, 95% CI [0.48, 1.46], z = −0.64, P = .523).

**Graph 4.**
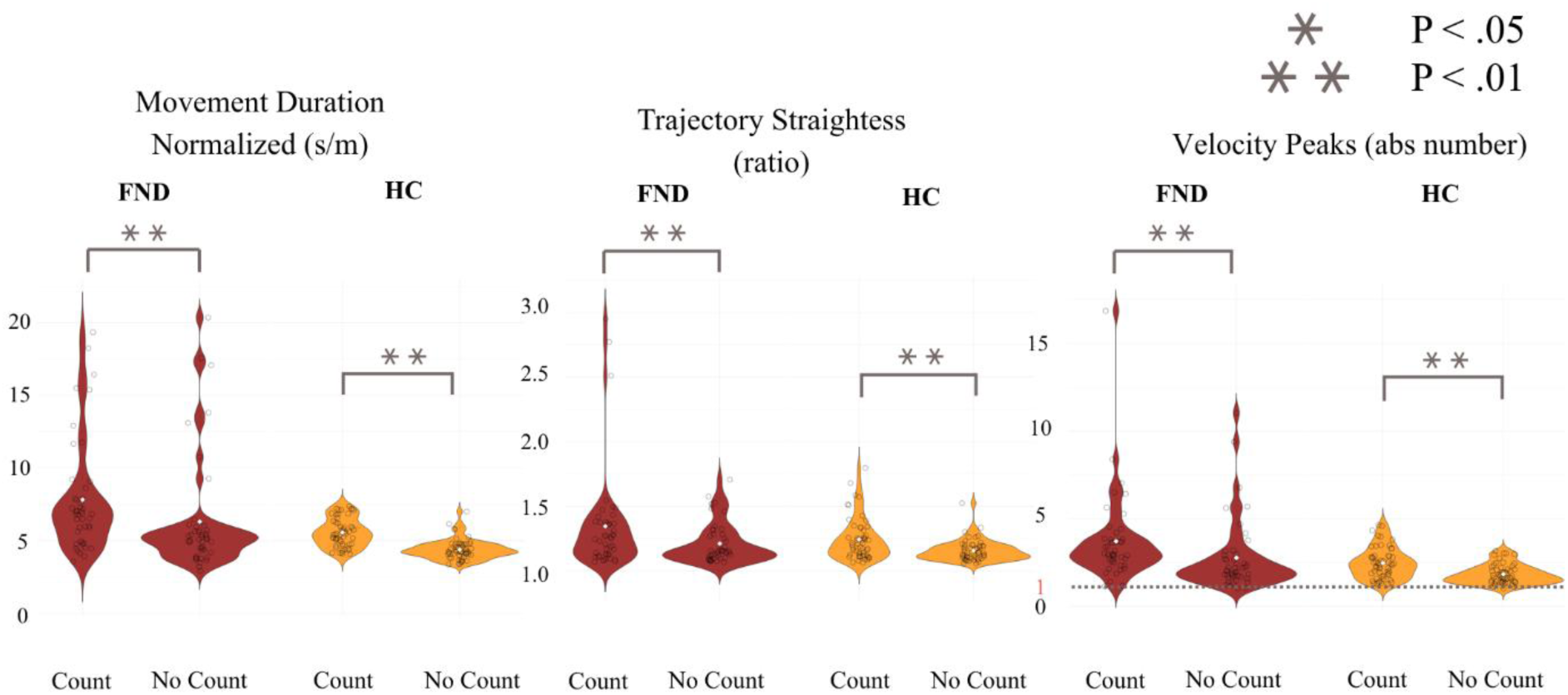
Representation of all modalities in both with and without the cognitive task. All the modalities have been merged to obtain only the effect of counting the fruits.

**Table 3.** Mean of each outcome by group and cognitive task.

| Participant Type | Cognitive task | Movement duration [mean (SD)] (s/m) | Straightness [mean (SD)] (ratio) | Number of peaks [mean (SD)] (absolute number) |
| --- | --- | --- | --- | --- |
| **Functional Neurological Disorder** | Count | 7.88 (5.24) | 1.33 (0.49) | 3.71 (4.28) |
|  | No count | 6.26 (6.26) | 1.21 (0.27) | 2.77 (3.47) |
| **Healthy Controls** | Count | 5.52 (1.59) | 1.22 (0.23) | 2.48 (1.95) |
|  | No count | 4.39 (1.18) | 1.15 (0.15) | 1.84 (1.37) |

### FND patients look more at their affected than unaffected arm

Within the AR condition, two of the four body regions (right arm and left arm) received no fixations in any trial and could not be analysed. For the right hand, the odds of body-directed gaze did not differ significantly between groups: relative to FND participants, HC showed lower but non-significant odds of fixation (b = −0.51, SE = 0.45, z = −1.14, p = .257; OR = 0.60). The same pattern held for the left hand (b = −0.69, SE = 0.58, z = −1.19, p = .233; OR = 0.50). Overall, FND patients looked at their right hand in 3.50% of trials, and HC 1.90%. For the left hand, FND looked in 1.93% of trials and HC during 1.23%. **Graph 5.**

**Graph 5.**
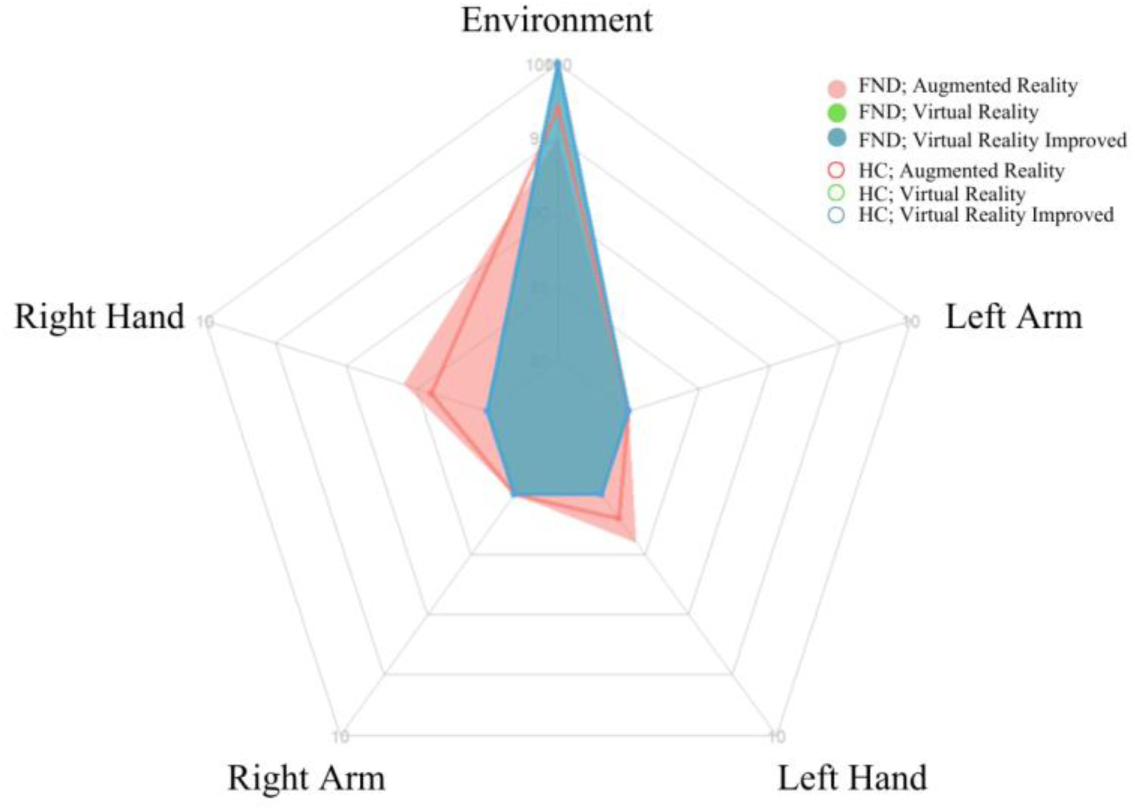
Representation of the proportions of trials in which participants looked at the targets. The first value at the center of the radar plot correspond to 0 trial with a fixation.

Within FND participants only, gaze allocation differed by limb affection for the right hand: the odds of fixating were significantly lower when it was the unaffected arm than the affected arm (b = −1.46, SE = 0.34, z = −4.23, p < .001; OR = 0.23), indicating that patients directed gaze toward their affected limb roughly four times more often. For the left hand, the effect was in the same direction but did not reach significance (b = −0.37, SE = 0.40, z = −0.92, p = .36; OR = 0.69).

### The repetition shows effects in the number of peaks for FND

Analyses comparing the first and last trials revealed no significant improvement in movement duration in any modality (AR: *b* = 0.427, *SE* = 0.392, *df* = 1005, *t* = 1.08, *p* = .276; VR: *b* = 0.157, *SE* = 0.379, *df* = 1005, *t* = 0.41, *p* = .678; VRI: *b* = −0.463, *SE* = 0.382, *df* = 1005, *t* = −1.21, *p* = .226). Similarly, movement straightness did not improve across modalities (AR: *b* = −0.015, *SE* = 0.047, *df* = 1002, *t* = −0.32, *p* = .749; VR: *b* = −0.049, *SE* = 0.046, *df* = 1002, *t* = −1.06, *p* = .287; VRI: *b* = 0.009, *SE* = 0.046, *df* = 1002, *t* = 0.21, *p* = .830).

In contrast, patients showed a significant reduction in the number of movement peaks across all modalities, indicating smoother trajectories over time (AR: *b* = −5.40, *SE* = 0.591, *df* = 1060, *t* = −9.12, *p* < .001; VR: *b* = −3.12, *SE* = 0.591, *df* = 1060, *t* = −5.27, *p* < .001; VRI: *b* = −3.63, *SE* = 0.591, *df* = 1060, *t* = −6.13, *p* < .001). **Graph 6.**

**Graph 6.**
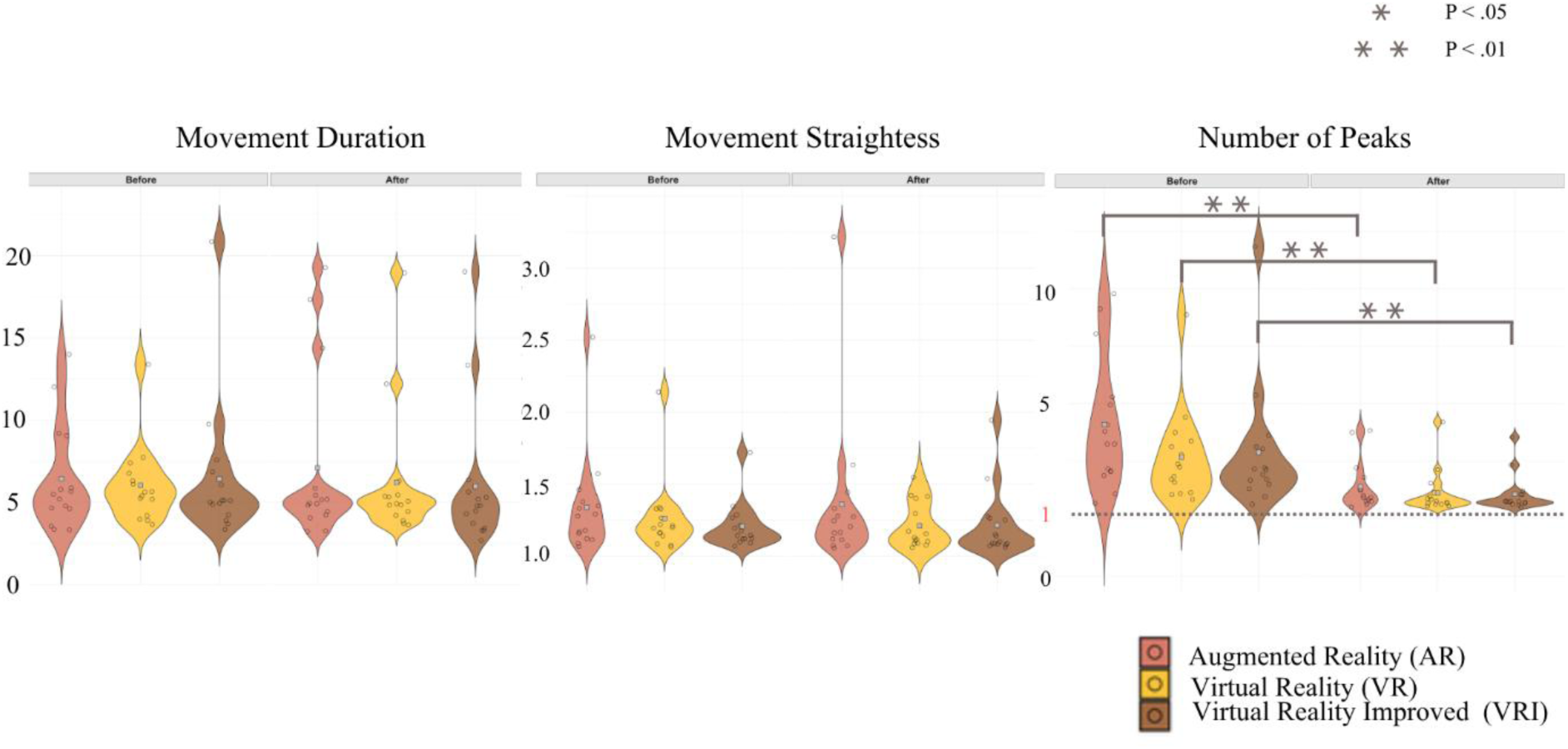
Comparison before and after training in FND. In all the three outcomes, only the number of Peaks has improved after training. All three modalities are significant with AR showing the most progress.

**Table 4.** Mean of each outcome by timepoint and modality.

| Modality | Timepoint | Movement duration<br>[mean (SD)] (s/m) | Straightness [mean<br>(SD)] (ratio) | Number of peaks<br>[mean (SD)]<br>(absolute number) |
| --- | --- | --- | --- | --- |
| **Augmented Reality (AR)** | Before | 6.43 (4.73) | 1.35 (0.45) | 8.2 (9.1) |
|  | After | 6.92 (6.60) | 1.33 (0.89) | 2.8 (3.45) |
| **Virtual Reality (VR)** | Before | 5.9 (2.65) | 1.25 (0.32) | 5.36 (9.24) |
|  | After | 6.15 (2.65) | 1.21 (0.45) | 2.24 (2.7) |
| **Virtual Reality Improved (VRI)** | Before | 6.35 (4.41) | 1.20 (0.21) | 5.76 (7.07) |
|  | After | 5.94 (4.77) | 1.21 (0.33) | 2.13 (1.92) |

### Physical reaching

Trials from the physical condition, in which participants reached for a real plastic ball without the head-mounted display, were not analysed. A fault in the recording setup left a high proportion of these trials unrecorded (logged as missing values), which precluded robust analysis. No trials were missing in the AR, VR or VRI conditions.

## Discussion

In this study, we investigated the effect of three different VR-based modalities on movement quality in FND patients and healthy controls. The result showed that FND participants were slower in the AR modality, when they see their arm, with HC showing the reverse pattern. Additionally, we also studied the impact of a dual motor-cognitive task and the effect of training by comparing the beginning and the end of the trial block for each modality, which shown only an improvement in the number of peaks.

During this experiment, all participants have completed the protocol as planned, showing a good feasibility.

### FND participants are similar to healthy controls when they do not see their arm

Focusing on the movement speed and smoothness, FND have shown to be slower with more adjustments in the Augmented Reality than in the two other modalities, although HC had an opposite pattern. A plausible interpretation for these opposite patterns is that the visibility of one’s real limb carries completely different cognitive and sensorimotor implications for FND patients and healthy controls. For healthy participants, seeing their own arm typically facilitates performance. For FND patients, however, the same visual feedback appears to have the opposite effect, due to pathological visuomotor integration. Indeed, motor symptoms of FND are thought to arise from an altered integration of feedforward and feedback signals, which might result in an imbalance between automatic motor control and attention-driven control. Thus, the AR condition, by making the patients’ real limb visible, can trigger excessive self-monitoring, increased attentional focus on the symptomatic limb, and heightened awareness of perceived “errors.” The result is slower, less fluid motor execution. This interpretation aligns with predictive-processing accounts of FND:^19^ when patients hold strong negative priors about their motor ability (“my movement will be wrong”), seeing their own limb may amplify the mismatch between expected and observed movement, leading them to consciously correct or inhibit their action. In VR and VRI, where the limb is replaced by a virtual avatar, FND participants are less anchored to their symptomatic body representation, reducing the amount of maladaptive self-focus. This allows movements to become more automatic, explaining why their performance improves relative to AR.

The movement straightness did not differ between groups and was always better in VR-Improved. This result appears even tough the reported data are the actual participant’s movement and not the improved avatar. This is probably due to the fact that visual feedback was enhanced, and participants were able to follow the path without readjusting the movement. This ended in a straighter movement.

### The cognitive task does not improve the performance in FND

In contrast to our expectations, based on the relief of symptoms while distracted,^4^ the additional cognitive task (i.e., counting fruits) did not improve motor performance in FND. Instead, FND participants were more negatively affected than healthy controls across all outcomes. Counting fruit within a virtual environment, however, does not reflect this usual form of distractibility and may have introduced an additional cognitive load rather than a beneficial distraction. Also, performing a dual cognitive-motor task could have different effects than an additive motor task and result in a worsening of symptoms instead of an improvement.

Current evidence regarding neurocognitive preservation in FND is mixed, with studies reporting inconsistent findings. Some work suggests cognitive deficits in domains such as working memory and cognitive flexibility. For example, Věchetová et al.^20^ found significantly poorer performance in FMD compared to healthy controls whereas other studies report no such differences.^21^ In our sample, the two groups did not differ in their counting accuracy, suggesting comparable task comprehension and basic cognitive performance. Nevertheless, the cognitive task had a disproportionately larger impact on motor performance in FND, indicating that even when accuracy is preserved, cognitive load may interfere more strongly with motor execution in this population.

### FND have more visual monitoring of their affected arm than unaffected arm

Our eye-tracking results did not reveal a significant difference in body-directed gaze between FND participants and healthy controls. Within FND patients, however, gaze allocation was biased by symptom laterality: participants looked significantly more at their affected limb than their unaffected one. This within-patient pattern suggests that attentional monitoring is preferentially directed toward the symptomatic limb rather than reflecting a generalised increase in self-directed attention distinguishing patients from controls. Previous accounts of heightened self-monitoring in FND have rested largely on indirect neuroimaging evidence;^22,23^ the present data provide a direct, behavioural correlate, though one specific to the affected limb rather than to the group as a whole.

From a mechanistic perspective, a laterality-specific attentional bias aligns with predictive-processing accounts of FND, which propose that strong, symptom-related priors (e.g., expectations of movement failure) dominate over sensory evidence. Visual attention plays a key role in this process: directing gaze toward a body part increases the precision, and hence the influence, assigned to predictions about it. Preferential monitoring of the affected limb may therefore amplify maladaptive expectations for that limb specifically, making its movements feel effortful or unreliable. That this bias emerged in AR, where the limb was clearly visible, is consistent with the idea that salient visual input can paradoxically strengthen prior-driven interpretation rather than correct it.

These findings complement early evidence of altered oculomotor control in FND, including abnormal saccades.^24,25^ Together, they suggest that visual monitoring and precision-weighting mechanisms may be disrupted in FND, potentially contributing to symptom maintenance. Further work will be required to determine whether this attentional bias represents a causal factor in symptom expression or an adaptive response to impaired movement.

### All modalities show an effect on movement smoothness, but not on speed not straightness

FND participants showed improvement in the number of velocity peaks across all modalities, suggesting that with repetition they required fewer corrective sub-movements to complete the task. Importantly, this did not translate into faster nor straighter movement duration, indicating that training helped them smooth their trajectories without increasing overall speed. All modalities showed some degree of improvement, with the largest gains appearing in AR, likely because this was initially the most challenging condition for FND. Interestingly, after training, their performance converged across modalities, suggesting that the negative impact of visual feedback can partially be reduced through repeated exposure. These findings imply that therapeutic interventions may benefit from focusing specifically on AR environments, where patients receive the most direct and ecologically valid feedback about their limb position. Future studies should incorporate longer training protocols and compare performance with standard motor rehabilitation tasks to determine whether these improvements generalize beyond the experimental setting.

### Limitations

This study has several limitations. First, technical issues during data collection led to the exclusion of a limited number of participants, potentially reducing the sensitivity of our analyses. Second, FND presentations are highly heterogeneous, yet our sample size did not allow for sub-group analyses (e.g., tremor vs. weakness, functional overlays, chronicity), limiting our ability to capture subtype-specific nuances. Additionally, the novelty of the task and the artificial laboratory environment may not fully reflect patients’ usual attentional strategies in clinical or daily-life contexts. Furthermore, because our study relied on cross-sectional behavioural data, we cannot determine whether the observed attentional and motor patterns reflect causal mechanisms or compensatory responses.

Concerning the physical reaching data, the transcription of these showed a low quality, with an important number of trials that weren’t correctly encoded. Those errors were checked and not present in the virtual data. Consequently, we decided to get rid of this part of the project.

Finally, concerning the training analysis, participants were only able to train for a limited amount of time and those results cannot translate to a more symptoms-focused intervention that would last for multiple days or weeks with repetitive training sessions.

### Conclusion

The present study investigated movement quality and visual behaviour across three technology-based modalities in individuals with FND compared with healthy controls. Our findings show that FND participants move significantly slower and less precisely, particularly when visual feedback of their body is available, and that they tend to monitor their own movements more extensively through increased gaze directed toward their affected limb.

This work also highlights the potential of immersive technologies to reveal symptom-relevant mechanisms in a controlled and, in future designs, ecologically valid environment. Additionally, the results of this study highlight that a VR setting - where participants see an avatar of their arm and tend to reduce their maladaptive self-monitoring - might be more beneficial than an AR setting where participants see their real arm. Future studies should examine whether repeated exposure, longer training protocols, or modality-specific interventions in virtual reality could modulate these attentional patterns and ultimately improve motor outcomes. Integrating clinical measures, longitudinal designs, and larger samples will be essential to develop specific VR-based therapeutic approaches as a complement to current rehabilitation strategies for FND.

## Supporting information

Supplementary results

## Data Availability

All data produced in the present study are available upon reasonable request to the authors

## Acknowledgements

We thank all the individuals who took part in this study for their valuable participation and trust. We also thank Manuela Steinauer for her active participation in the project. We also thank Michaël Mouthon for his help in the data analysis.

## Supplementary material

[ARVR_SupplementaryResults](https://docs.google.com/document/d/1BPYn9bK9a06rVJHIk4IjRdXqlBgJOk-S4kqayL1a2PY/edit?tab=t.0)

