## Supplementary results for "Virtual environment benefits motor dysfunction in Functional Neurological Disorder: Evidence from Virtual and Augmented Reality"

Full address: Ch. du Musée 5, 1700 Fribourg

### The modality mostly affected questionnaires, but not participant type

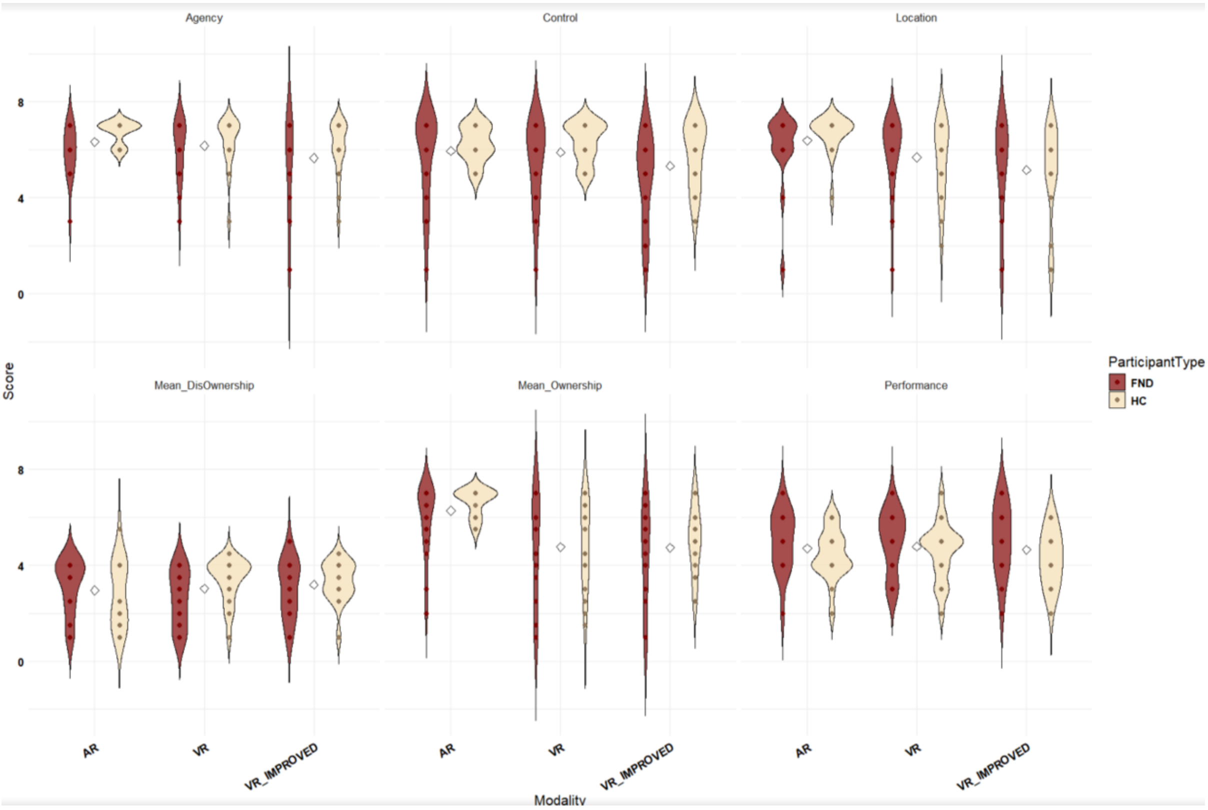


**Supplementary Figure 1.** Distribution of scores for each questionnaire scale across all modalities (AR, VR, VRI), shown separately for FND patients and healthy controls

| Questionnaire Scale | Effect | F(DFn, DFd) | p-value |
|---|---|---|---|
| **Agency** | Participant Type | 7.34 (1, 30) | 0.011* |
| | Modality | 3.38 (2, 60) | 0.041* |
| | Participant Type × Modality | 0.54 (2, 60) | 0.587 |

| | | | |
|---|---|---|---|
| **Control** | Participant Type | 3.13 (1, 30) | 0.087 |
| | Modality | 6.32 (2, 60) | 0.003* |
| | Participant Type × Modality | 0.64 (2, 60) | 0.529 |
| **Location** | Participant Type | 0.10 (1, 29) | 0.752 |
| | Modality | 6.03 (2, 58) | 0.004* |
| | Participant Type × Modality | 0.97 (2, 58) | 0.387 |
| **Mean_Ownership** | Participant Type | 1.11 (1, 30) | 0.301 |
| | Modality | 14.50 (2, 60) | 7.30e-06* |
| | Participant Type × Modality | 1.09 (2, 60) | 0.343 |
| **Mean_DisOwnership** | Participant Type | 1.33 (1, 30) | 0.258 |
| | Modality | 0.70 (2, 60) | 0.500 |
| | Participant Type × Modality | 3.15 (2, 60) | 0.050 |
| **Performance** | Participant Type | 2.76 (1, 30) | 0.107 |
| | Modality | 0.31 (2, 60) | 0.735 |
| | Participant Type × Modality | 0.08 (2, 60) | 0.926 |

**Supplementary Table 1.** Summary of main effects and interactions from the mixed ANOVA (Participant Type × Modality) for each questionnaire scale. Asterisks denote $p < .05$.

### FND patients are more impacted by the cognitive task in all modalities

Augmented reality - Movement Duration normalized

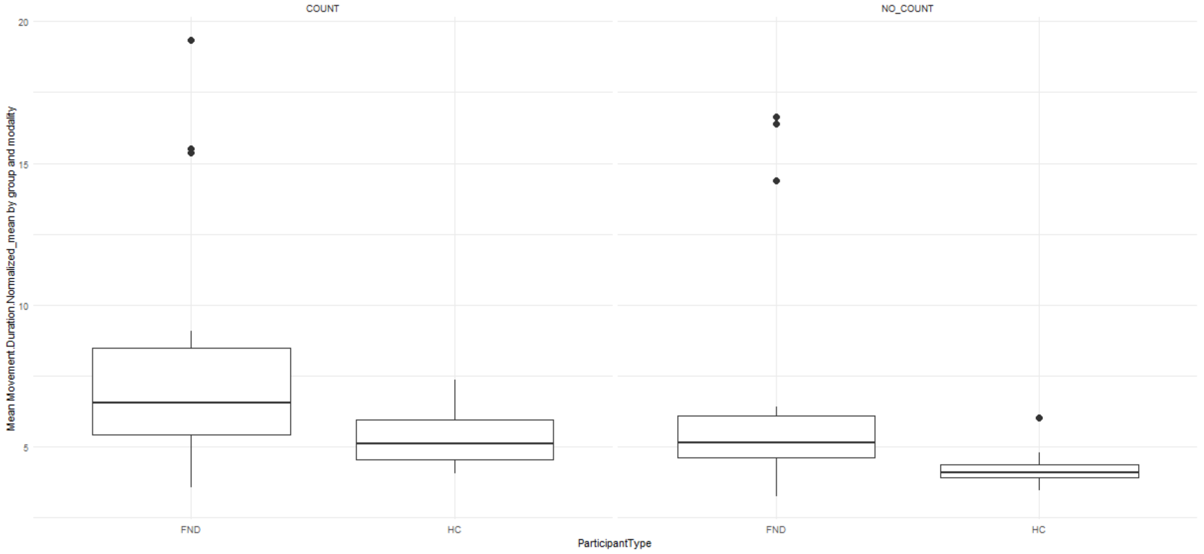


Post-hoc contrasts showed a significant effect of modality in both groups (FND: est. = 1.16, SE = 0.18, $t(2808.5) = 6.42$, $p < .001$; HC: est. = 1.09, SE = 0.17, $t(2808.5) = 6.55$, $p < .001$), with higher values in COUNT than NO_COUNT. FND patients scored higher than HC in both modalities (COUNT: est. = 2.90, SE = 1.11, $t(32.3) = 2.60$, $p = .014$; NO_COUNT: est. = 2.83, SE = 1.11, $t(31.4) = 2.56$, $p = .016$).

Augmented reality - Movement Straightness

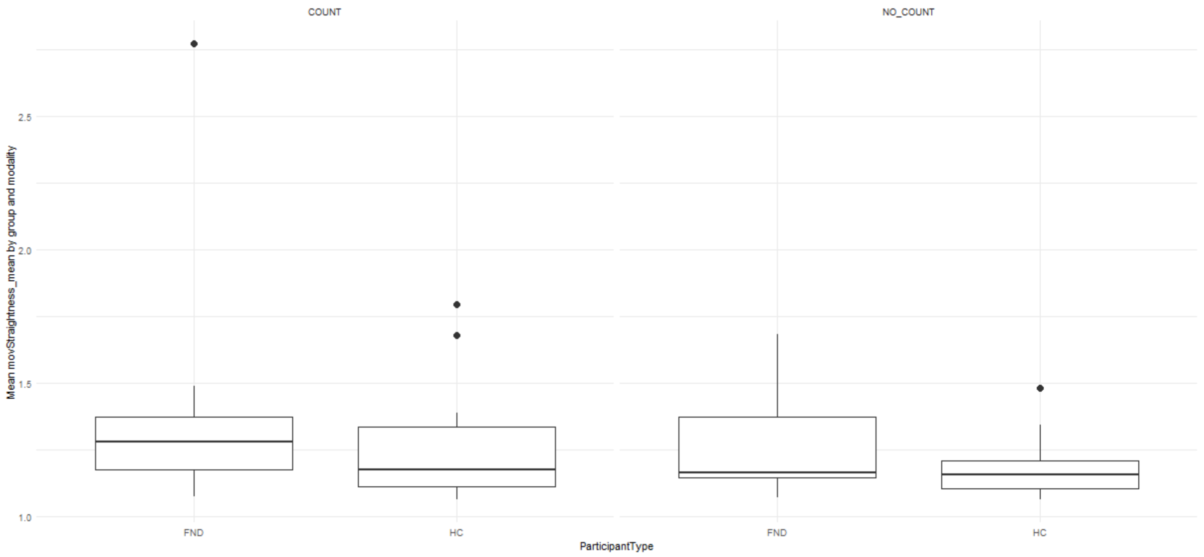


For movement straightness, the COUNT–NO_COUNT contrast was significant in both groups (FND: est. = 0.096, SE = 0.018, $t(2819.3) = 5.22$, $p < .001$; HC: est. = 0.064, SE = 0.017, $t(2819.4) = 3.73$, $p$

< .001). Group differences were not significant in either modality (COUNT: est. = 0.121, SE = 0.071, $t(34.6) = 1.71$, $p = .096$; NO_COUNT: est. = 0.089, SE = 0.069, $t(32.1) = 1.28$, $p = .209$).

Augmented reality - Number of Peaks

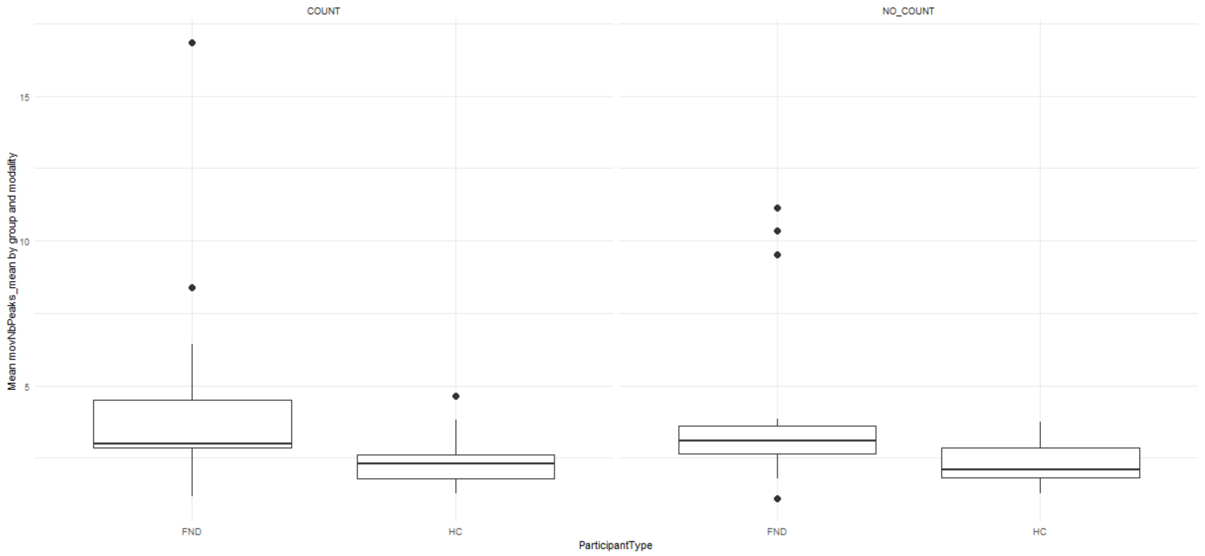


For the number of movement peaks, the COUNT–NO_COUNT contrast was not significant in either group (FND: est. = 0.275, SE = 0.208, $z = 1.32$, $p = .186$; HC: est. = 0.024, SE = 0.190, $z = 0.13$, $p = .899$). FND patients showed significantly more peaks than HC in both modalities (COUNT: est. = 2.22, SE = 0.81, $z = 2.73$, $p = .006$; NO_COUNT: est. = 1.97, SE = 0.80, $z = 2.46$, $p = .014$).

Virtual reality - Movement Duration normalized

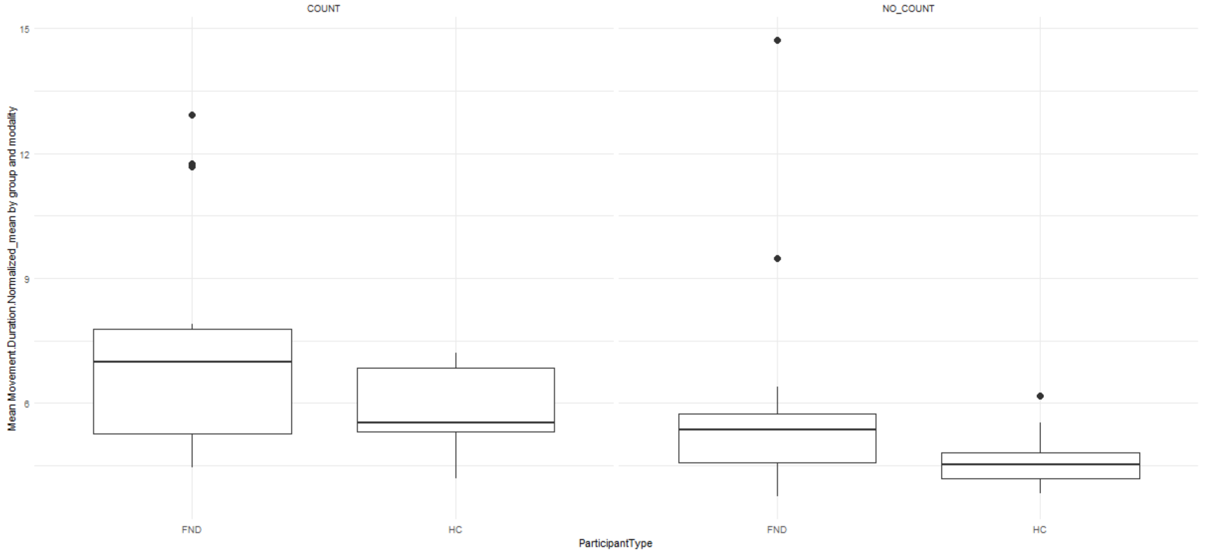


For normalized movement duration, the COUNT–NO_COUNT contrast was highly significant in both groups (FND: est. = 1.368, SE = 0.109, $t(2916.3) = 12.53$, $p < .001$; HC: est. = 1.189, SE = 0.102, $t(2916.6) = 11.61$, $p < .001$). FND patients showed longer normalized durations than HC in both modalities (COUNT: est. = 1.543, SE = 0.639, $t(32.5) = 2.41$, $p = .022$; NO_COUNT: est. = 1.364, SE = 0.634, $t(31.4) = 2.15$, $p = .039$).

Virtual reality - Movement Straightness

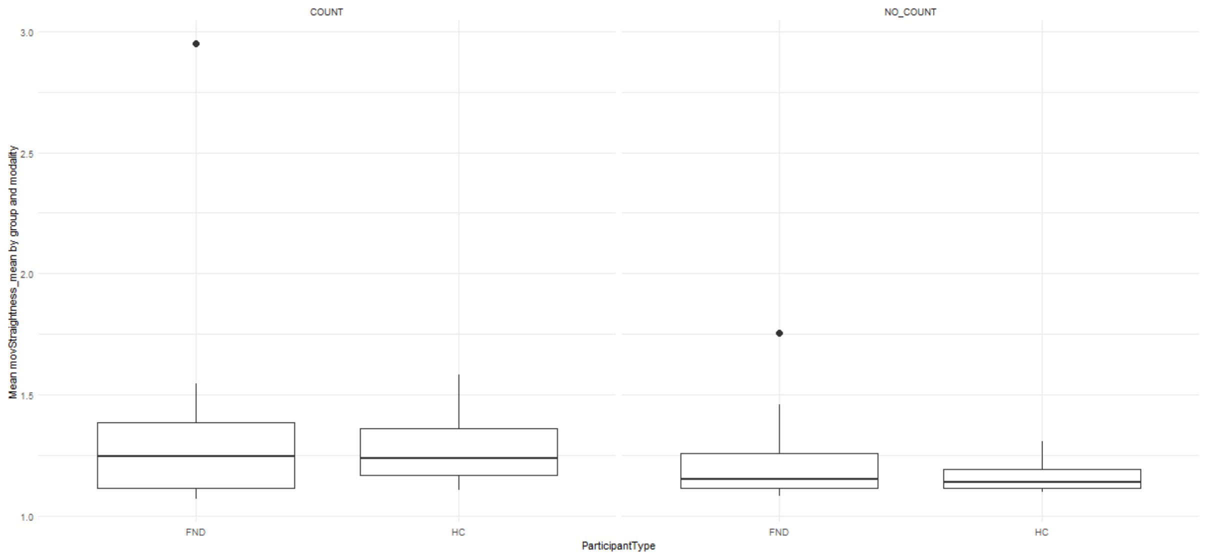


For movement straightness, the COUNT–NO_COUNT contrast was significant in both groups (FND: est. = 0.129, SE = 0.014, $t(2961.1) = 9.56$, $p < .001$; HC: est. = 0.108, SE = 0.013, $t(2963.2) = 8.33$, $p < .001$). Group differences were not significant in either modality (COUNT: est. = 0.078, SE = 0.069, $t(33.0) = 1.13$, $p = .268$; NO_COUNT: est. = 0.057, SE = 0.069, $t(31.6) = 0.83$, $p = .411$).

Virtual reality - Number of Peaks

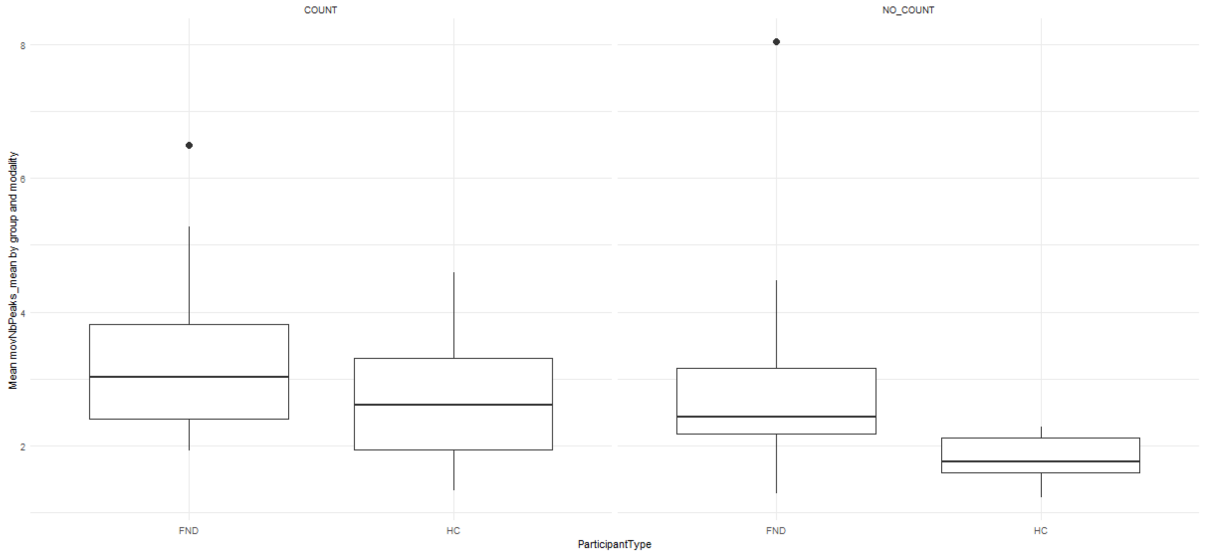


For the number of movement peaks, the COUNT–NO_COUNT contrast was significant in both groups (FND: est. = 0.350, SE = 0.161, $z = 2.17$, $p = .030$; HC: est. = 0.889, SE = 0.147, $z = 6.04$, $p < .001$). FND patients showed significantly more peaks than HC in the NO_COUNT condition (est. = 1.143, SE = 0.339, $z = 3.37$, $p < .001$), whereas the group difference did not reach significance under COUNT (est. = 0.604, SE = 0.356, $z = 1.70$, $p = .090$).

Virtual reality augmented - Movement Duration normalized

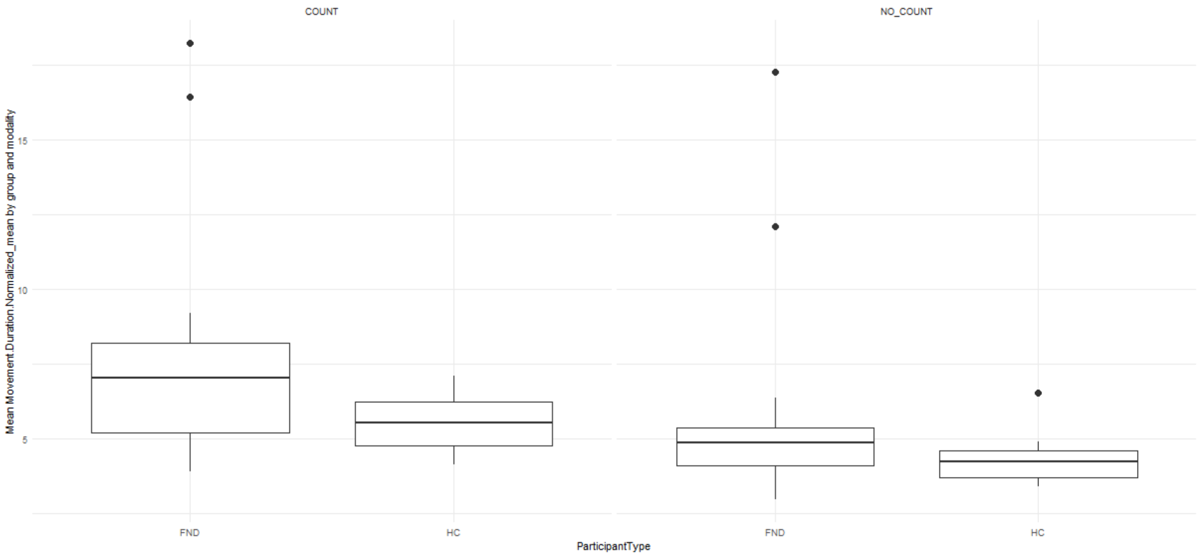


For normalized movement duration, the COUNT–NO_COUNT contrast was highly significant in both groups (FND: est. = 1.885, SE = 0.118, $z = 15.94$, $p < .001$; HC: est. = 1.220, SE = 0.107, $z = 11.40$, $p < .001$). FND patients showed significantly longer normalized durations than HC under COUNT (est. = 2.312, SE = 0.939, $z = 2.46$, $p = .014$), whereas the group difference did not reach significance under NO_COUNT (est. = 1.647, SE = 0.935, $z = 1.76$, $p = .078$).

Virtual reality augmented - Movement Straightness

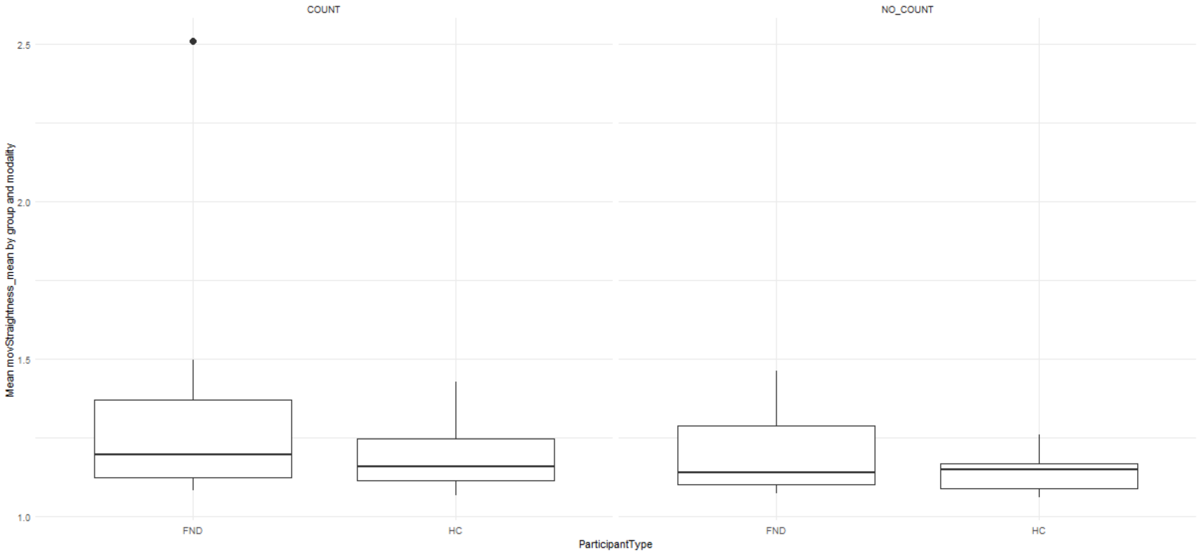


For movement straightness, the COUNT–NO_COUNT contrast was significant in both groups (FND: est. = 0.101, SE = 0.012, $z = 8.24$, $p < .001$; HC: est. = 0.049, SE = 0.011, $z = 4.34$, $p < .001$). FND patients differed significantly from HC under COUNT (est. = 0.109, SE = 0.050, $z = 2.18$, $p = .030$), whereas the group difference was not significant under NO_COUNT (est. = 0.056, SE = 0.049, $z = 1.14$, $p = .254$).

Virtual reality augmented - Number of Peaks

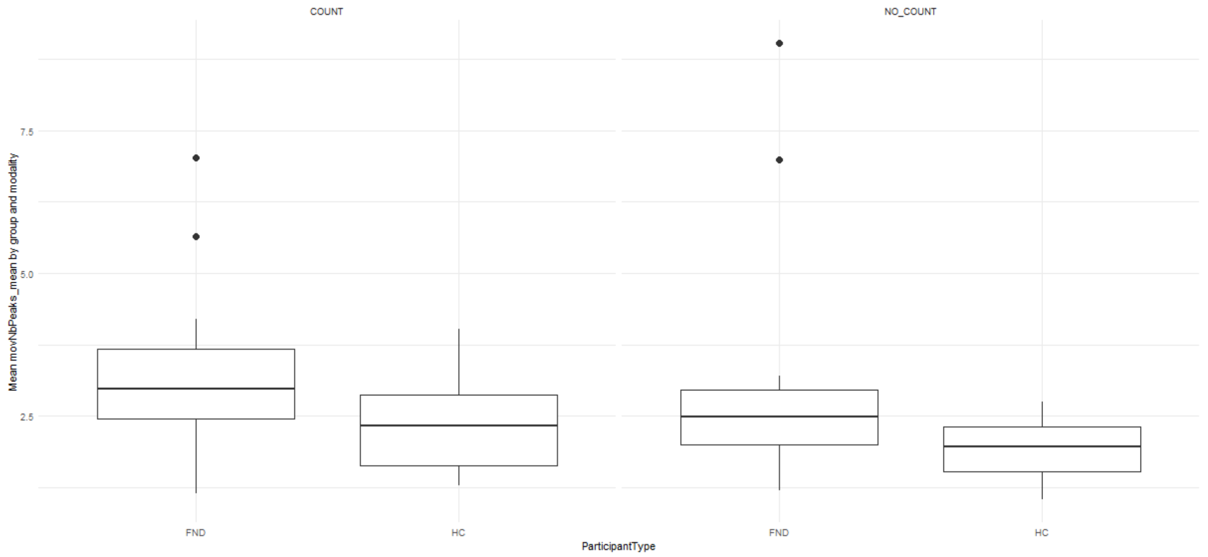


For the number of movement peaks, the COUNT–NO_COUNT contrast was significant in HC (est. = 0.448, SE = 0.120, $z$ = 3.72, $p$ < .001) but not in FND (est. = 0.227, SE = 0.132, $z$ = 1.72, $p$ = .085). FND patients showed significantly more peaks than HC under NO_COUNT (est. = 1.106, SE = 0.461, $z$ = 2.40, $p$ = .016), with a marginal difference under COUNT (est. = 0.886, SE = 0.470, $z$ = 1.89, $p$ = .059).
